# Urban pandemic response: survey results describing the experiences from twenty-five cities during the COVID-19 pandemic

**DOI:** 10.1101/2022.07.12.22277559

**Authors:** Matthew R Boyce, Melissa Cordoba Asprilla, Breanna van Loenen, Amanda McClelland, Ariella Rojhani

## Abstract

Since first being detected in Wuhan, China in late December 2019, the COVID-19 pandemic has demanded a response from all levels of government. While the role of local governments in routine public health functions is well understood, and the response to the pandemic has highlighted the importance of involving local governments in the response to and management of large, multifaceted challenges, their role in pandemic response remains more opaque. Accordingly, to better understand how local governments in cities were involved in the response to the COVID-19 pandemic, we conducted a survey involving cities in the Partnership for Healthy Cities to: (i) understand which levels of government were responsible, accountable, consulted, and informed regarding select pandemic response activities; (ii) document when response activities were implemented; (iii) characterize how challenging response activities were; and (iv) query about future engagement in pandemic and epidemic preparedness. Twenty-five cities from around the world completed the survey and we used descriptive statistics to summarize the urban experience in pandemic response. Our results show that national authorities were responsible and accountable for a majority of the activities considered, but that local governments were also responsible and accountable for key activities – especially risk communication and coordinating with community-based organizations and civil society organizations. Further, most response activities were implemented after COVID-19 had been confirmed in a city, many pandemic response activities proved to be challenging for local authorities, and nearly all local authorities envisioned being more engaged in pandemic preparedness and response following the COVID-19 pandemic. This descriptive research represents an important contribution to an expanding evidence base focused on improving the response to the ongoing COVID-19 pandemic, as well as future outbreaks.

## Introduction

On December 31, 2019, the World Health Organization (WHO) China country office was notified about a series of pneumonia cases of unknown etiology in the city of Wuhan, Hubei Province [1]. Over the next several weeks, cases were confirmed in Thailand (January 12, 2020), Japan (January 15, 2020), South Korea (January 20, 2020), and the United States of America (January 21, 2020), all in travelers incoming from Wuhan [1]. On January 30, 2020, with hundreds of cases identified across 19 countries, the WHO declared the outbreak a Public Health Emergency of International Concern [2]. And on March 11, 2020, with over 118,000 cases of a disease now known as COVID-19 being reported across 114 countries, the WHO characterized the outbreak as a pandemic [3].

The significant societal risks posed by pandemics have long been recognized by the public health community. However, the rapid and global spread of COVID-19 caught the world by surprise; and governments, at all levels, have since grappled with the difficult tasks associated with mounting responses to combat the resulting health, social, and economic consequences. This includes local-level governments and authorities – especially those in cities. Because of their dense populations and high degree of connectivity, cities can represent a rate-enhancing or - limiting gateway for infectious disease transmission [4]. Further, neglecting local contexts and associated complexities can prove disastrous for response efforts, as it is the local level that most often responds first to events like infectious disease outbreaks [5]. As such, understanding the detection, response, and subsequent control of infectious disease outbreaks in cities represent key concerns for the public health and policy communities.

This is especially true for the response to the COVID-19 pandemic, where some have argued that overlooking local contexts may have important implications for explaining why certain responses have been perceived as relatively successful, while others have failed [6]. For example, despite the recognized importance of the local knowledge essential for contact tracing [7], contact tracing during the COVID-19 pandemic has frequently been outsourced to entities with little experience or local knowledge – ultimately hampering efforts to control the outbreak [6].

Others have discussed how the COVID-19 pandemic has highlighted the increasing importance of involving local governments in the management of complex challenges [8]. With regard to epidemics and pandemics, the extent to which a city is involved largely depends on the political context in which it exists and the level of independence it has in relation to national or subnational governments (i.e., state, provincial, district, etc.) [9-12]. For instance, municipal authorities with clear administrative and financial autonomy may be fully in charge of managing and implementing a public health response, whereas others may depend heavily upon assistance from other levels [10, 13].

However, while these aforementioned statements are widely accepted at a broad level, important questions remain about the details of such arrangements and how response activities are coordinated during the response to actual outbreaks. This is especially true for a pandemic response, which has led to tensions between levels of government because of the novel nature of the event [14]. Accordingly, to better understand how activities were implemented and coordinated in cities to respond to the COVID-19 pandemic, we conducted a survey of cities that are a part of the Partnership for Healthy Cities (PHC). The specific objectives of the study were to: (i) understand which levels of government were responsible, accountable, consulted, and informed regarding select pandemic response activities; (ii) document when response activities were implemented in cities; (iii) characterize how challenging various response activities were; and (iv) query about future engagement in pandemic and epidemic preparedness.

## Methods

### Study Population

The study population involved in this research included local public health authorities from cities that are members of the PHC. The PHC is a global network of cities that have committed to using evidence-based approaches and interventions to prevent noncommunicable diseases and injuries. In March of 2020, the network expanded its scope beyond noncommunicable diseases and injuries to temporarily include pandemic response. Fifty cities involved in the PHC were purposively selected for this study, based on their involvement in a program focused on bolstering the local response to the COVID-19 pandemic.

### Questionnaire Development and Distribution

We developed a survey questionnaire to characterize the coordination of select pandemic response activities, when activities were implemented, and how challenging activities were. The survey was comprised of 30 questions and contained questions in a variety of formats including free-response, Likert-scales, matrices, and multiple-choice questions. We used Responsible, Accountable, Consulted, and Informed (RACI) matrices – a project management tool that is useful for investigating coordination structures – to investigate the governance of pandemic response and how activities were coordinated in cities. The survey questionnaire is available for review in the S1 Appendix.

For the purposes of this study, we defined responsible as the level of government that implemented the work required to complete an activity; accountable was the level that oversaw the correct and thorough completion of an activity; consulted was defined as levels engaging in two-way communication to provide information necessary for the completion of an activity; and informed was defined as a level of government that was updated regarding a given activity (i.e., one-way communication) [15]. Only one level of government could be accountable for a given activity, but multiple levels could be responsible, consulted, and informed. Further, we defined the national level as relating to the efforts of the national government; the subnational level as relating to the efforts of state, provincial, district, or county governments; and the local level as relating to the efforts of city or municipal governments.

Once finalized, we translated the questionnaire and accompanying instructions into French and Spanish to improve the accessibility and comprehensibility of study materials for a multilingual study population. We then uploaded the questionnaire and instructions to Qualtrics (Seattle, WA) and distributed them to local-level authorities in June 2021. The Qualtrics survey link expired in August 2021.

### Key Informant Interviews

Following the completion of surveys, we validated the results by conducting key informant interviews with several study participants. We purposively selected seven cities to ensure contextual diversity (i.e., geographic, development/income, etc.). The seven cities selected were: Accra, Ghana; Addis Ababa, Ethiopia; Bandung, Indonesia; Bengaluru, India; Kampala, Uganda; Lima, Peru; and London, United Kingdom. All interviews were conducted over Zoom (San Jose, CA) between June 2021 and August 2021.

### Data Analysis

We collected data from survey responses and key informant interviews and compiled them in a Microsoft Excel spreadsheet (Redmond, WA). We reviewed data for consistency and validity before using Microsoft Excel and STATA v.17BE (College Station, TX) to perform basic descriptive statistical analyses.

### Ethics Statement

We submitted all study materials for ethical review to the Georgetown University Institutional Review Board, which determined that the research posed no more than minimal risk to study participants and granted an exemption to the study (STUDY00003948). Written or oral informed consent was, thus, not required. However, as a best practice, we provided an informed consent form to all study participants that emphasized the voluntary nature of their participation, that their participation was not incentivized, and that they maintained the option of withdrawing from the study at any time without consequence. Consent was then provided through participation in the study.

## Results

### Study Participants

Local-level health authorities from 25 cities – Accra, Ghana; Addis Ababa, Ethiopia; Amman, Jordan; Athens, Greece; Bandung, Indonesia; Bangkok, Thailand; Barcelona, Spain; Bengaluru, India; Buenos Aires, Argentina; Cali, Colombia; Colombo, Sri Lanka; Guadalajara, Mexico; Kampala, Uganda; Kigali, Rwanda; Kumasi, Ghana; Lima, Peru; London, United Kingdom; Lusaka, Zambia; Medellín, Colombia; Ouagadougou, Burkina Faso; Rio de Janeiro, Brazil; Santiago, Chile; Santo Domingo, Dominican Republic; Vancouver, Canada; and Yangon, Myanmar – completed surveys. One city did not provide complete responses to the portions relating to how challenging pandemic response was or future efforts and engagement in pandemic preparedness and response. Five (20.0%) participating cities were located in countries that are currently classified as high-income countries, 10 (40.0%) were located in upper-middle-income countries, six (24.0%) were located in lower-middle-income countries, and four (16.0%) were located in low-income countries (Table 1). A majority (60%) of study participants were female and most (56%) had worked in their current professional role for 1 to 4 years.

**Table 1.** Characteristics of study participants and cities (n = 25).

| Characteristic | n (%) |
| --- | --- |
| Gender of Study Participant |  |
| *Male* | 10 (40.0) |
| *Female* | 15 (60.0) |
| *Other/Prefer not to say* | 0 (0.0) |
| Number of Years Served in Current Role |  |
| *Less than 1 year* | 5 (20.0) |
| *1 to 4 years* | 14 (56.0) |
| *5 to 9 years* | 4 (16.0) |
| *10 years or more* | 2 (8.0) |
| Economy Income Level |  |
| *High income* | 5 (20.0) |
| *Upper-middle income* | 10 (40.0) |
| *Lower-middle income* | 6 (24.0) |
| *Low income* | 4 (16.0) |

### Pandemic Response Activities – Coordination, Timing, and Challenges

RACI matrices responses revealed various coordination structures for pandemic response activities. In some, such as those in Amman, Kumasi, and Santo Domingo, the national government maintained primary authority and was accountable for virtually all pandemic response activities in the city; in other cities, such as Buenos Aires, Santiago, and Vancouver, subnational governments maintained primary authority for response activities; and in other cities, such as Bengaluru, Kampala, and Medellín, the local governments maintained primary authority. However, most often, the accountability and responsibility for pandemic response activities were shared across the various levels of government. Reported RACI matrices are available for review in the S2 Appendix.

Relative to other levels of government, the national government was responsible for a majority of the activities considered (Fig 1). National authorities were most frequently reported to be responsible for 11 of the 16 pandemic preparedness activities, national and subnational authorities for two (i.e., maintaining essential health services and providing diagnostic testing services), subnational authorities for one (i.e., contact tracing activities), and local authorities for two of the 16 activities (i.e., risk communication activities and coordinating with community-based and civil society organizations).

**Fig 1.**
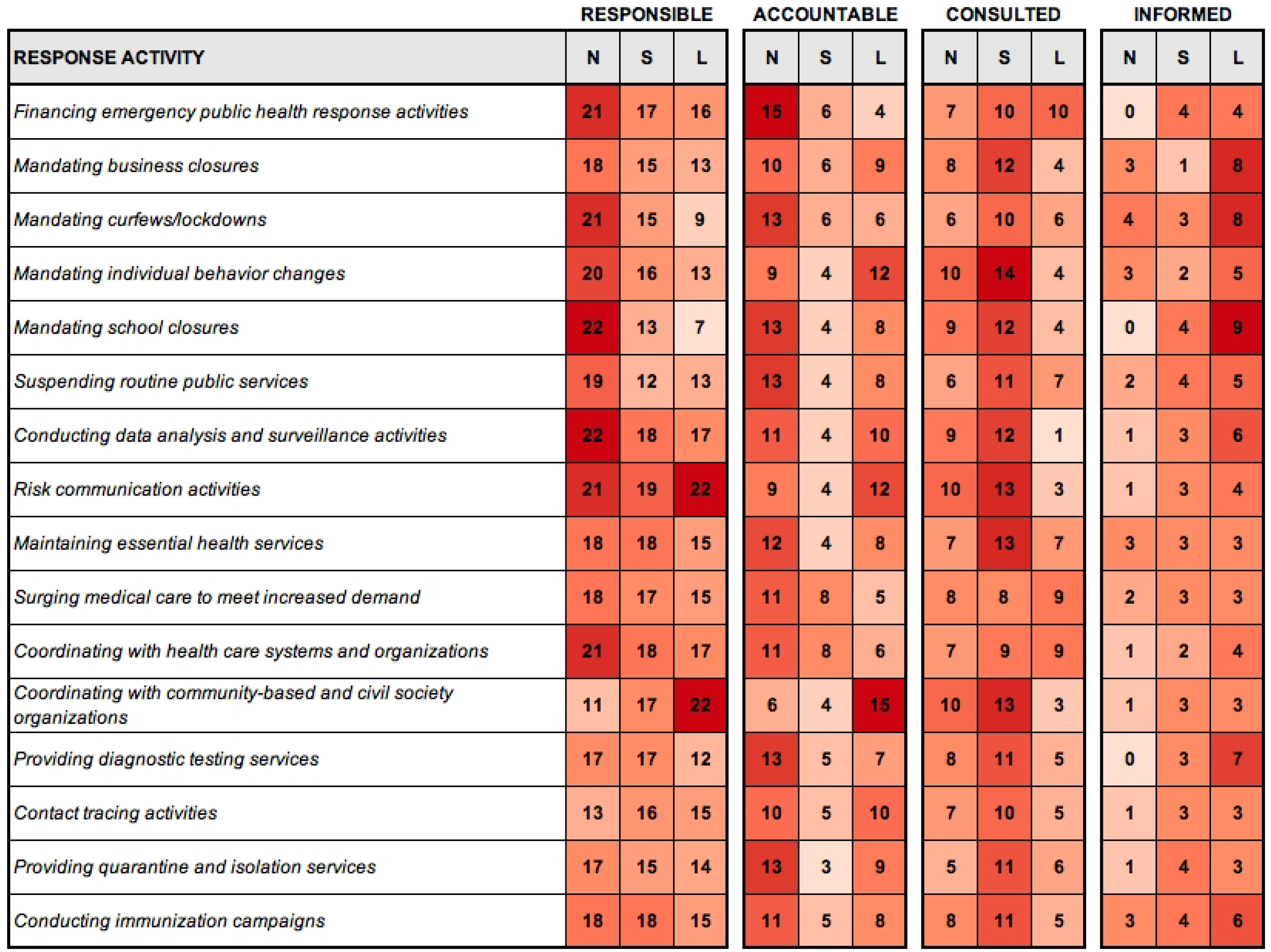
Heat maps displaying which levels of government are responsible, accountable, accounted, and informed for select pandemic response activities in cities according to survey responses (n = 25). One city indicated that no level of government was accountable for maintaining essential health services, surging medical care to meet increased demand, or conducting immunization campaigns; N – National, S – Subnational, L – Local.

Likewise, relative to other levels of government, the national level was most often accountable for 12 of the 16 activities, the national and local level for one activity (i.e., contact tracing activities), and the local level for three activities (i.e., mandating individual behavior changes, risk communication activities, and coordinating with community-based and civil society organizations). One city indicated that no level of government was accountable for maintaining essential health services, surging medical care to meet increased demand, or conducting immunization campaigns. Survey responses also demonstrated that different levels of government frequently communicated with one another – with those at the subnational level often consulted, and those at the local level often informed regarding response activities.

Survey responses indicated a relatively reactive response, as opposed to a proactive response. A majority of the cities surveyed indicated that the local government implemented activities after COVID-19 had been confirmed in the city for 11 of the 16 activities considered (i.e., mandating business closures, mandating curfews/lockdowns, mandating individual behavior changes, mandating school closures, surging medical care, coordinating with community-based and civil society organizations, providing diagnostic testing services, contact tracing activities, providing quarantine and isolation services, conducting immunization campaigns) (Table 2). The response activities that were most often implemented proactively (i.e., before COVID-19 had been confirmed in the country) were risk communication activities and coordinating with health care systems and organizations.

**Table 2.** Time that pandemic response activities were implemented by local governments (n = 25); Community-based organizations (CBOs), Civil society organizations (CSOs).

| Pandemic Response Activity | Time Response Activity was Implemented by Local Government (%) |  |  |  |
| --- | --- | --- | --- | --- |
|  | Before confirmed in country | Confirmed in country, not city | After confirmed in city | Not implemented or not applicable |
| Financing emergency public health response activities | 7 (28) | 8 (32) | 8 (32) | 2 (8) |
| Mandating business closures | 1 (4) | 3 (12) | 17 (68) | 4 (16) |
| Mandating curfews/lockdowns | 2 (8) | 3 (12) | 16 (64) | 4 (16) |
| Mandating individual behavior changes | 3 (12) | 7 (28) | 14 (56) | 1 (4) |
| Mandating school closures | 1 (4) | 5 (20) | 16 (64) | 3 (12) |
| Suspending routine public services | 0 (0) | 5 (20) | 17 (68) | 3 (12) |
| Conducting data analysis and surveillance activities | 8 (32) | 4 (16) | 11 (44) | 2 (8) |
| Risk communication activities | 10 (40) | 4 (16) | 9 (36) | 2 (8) |
| Maintaining essential health services | 10 (40) | 2 (8) | 11 (44) | 2 (8) |
| Surging medical care to meet increased demand | 4 (16) | 3 (12) | 15 (60) | 3 (12) |
| Coordinating with health care systems and organizations | 10 (40) | 4 (16) | 10 (40) | 1 (4) |
| Coordinating with CBOs and CSOs | 5 (20) | 5 (20) | 14 (56) | 1 (4) |
| Providing diagnostic testing services | 4 (16) | 3 (12) | 15 (60) | 3 (12) |
| Contact tracing activities | 2 (8) | 2 (8) | 18 (72) | 3 (12) |
| Providing quarantine and isolation services | 3 (12) | 2 (8) | 17 (68) | 3 (12) |
| Conducting immunization campaigns | 1 (4) | 3 (12) | 16 (64) | 5 (20) |

Survey responses also showed that many of the response activities were viewed as challenging. A minimum of half of the study participants described financing emergency public health response activities, mandating business closures, mandating curfews/lockdowns, mandating individual behavior changes, suspending routine public services, maintaining essential health services, surging medical care, and providing quarantine and isolation services as either extremely challenging or very challenging (Fig 2). Mandating business closures and mandating individual behavior changes (i.e., facemasks, social distancing, etc.) were reportedly the most challenging response activities, with 12 participants (50.0%) characterizing these activities as extremely challenging and another six (25.0%) characterizing them as very challenging. Financing emergency public health response activities was another reportedly challenging activity, with five participants (20.8%) characterizing this activity as extremely challenging and another 13 (54.2%) characterizing it as very challenging. Conducting surveillance & data analysis activities was the response activity characterized as the least challenging, relative to others considered in this study; four participants (16.7%) characterized it as not challenging and another four (16.7%) characterized it as slightly challenging.

**Fig 2.**
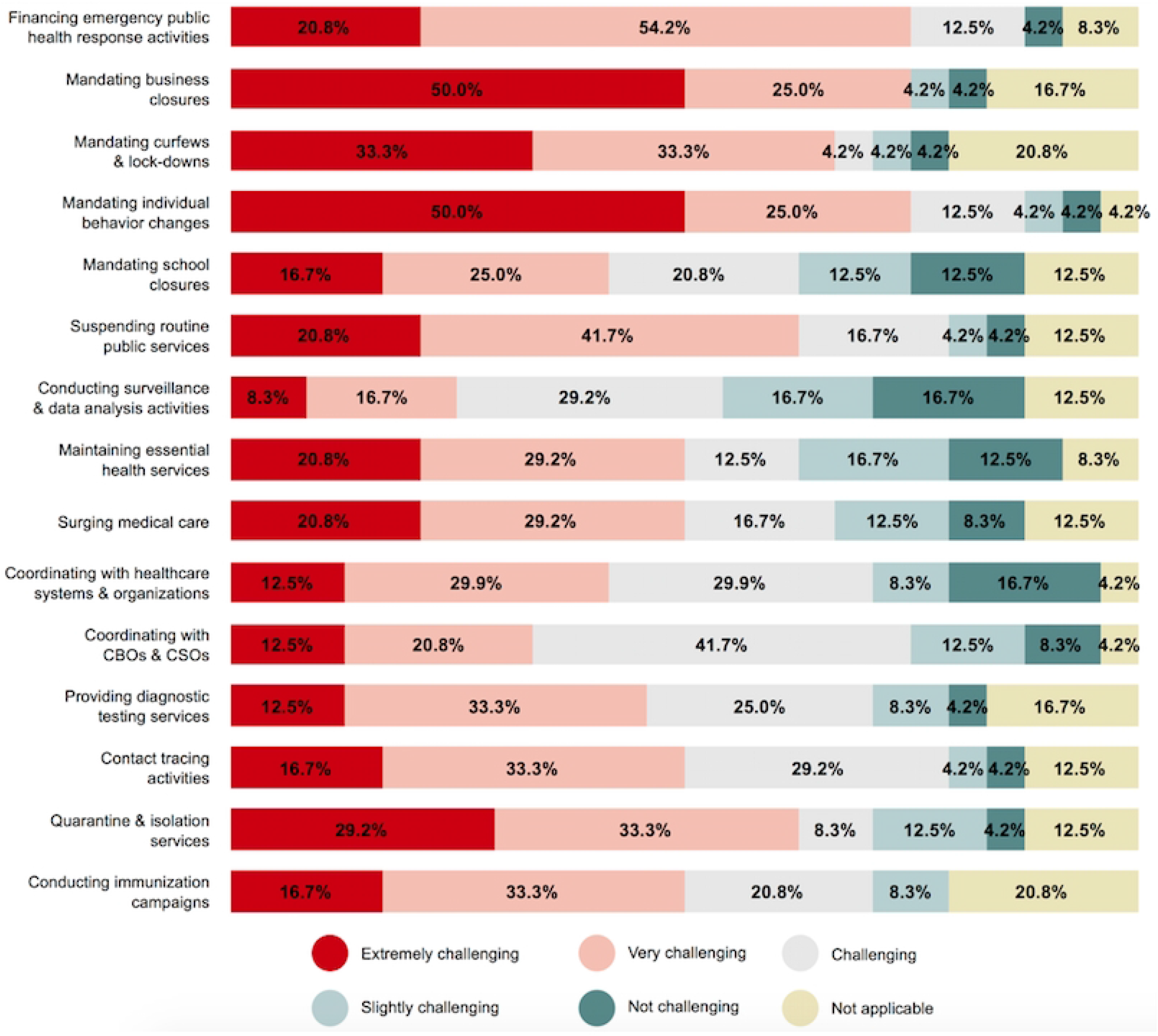
Likert-scale results regarding how challenging pandemic response activities were perceived to be in cities (n = 24); Community-based organizations (CBOs), Civil society organizations (CSOs).

When asked about specific risk communication activities, survey responses showed that reaching high-risk populations with information and addressing misinformation and disinformation were the two most challenging activities, with at least half of study participants characterizing these activities as either extremely challenging or very challenging (Fig 3). Addressing misinformation and disinformation proved to be the most challenging of the communication activities asked about, with nine (37.5%) of the study participants characterizing it as extremely challenging for their city and another 10 (41.7%) characterizing it as very challenging. Internal communications (i.e., within the local government) was the least challenging communication activity, with six (25.0%) of the study participants characterizing it as not challenging for their city and another two (8.3%) characterizing it as only slightly challenging.

**Fig 3.**
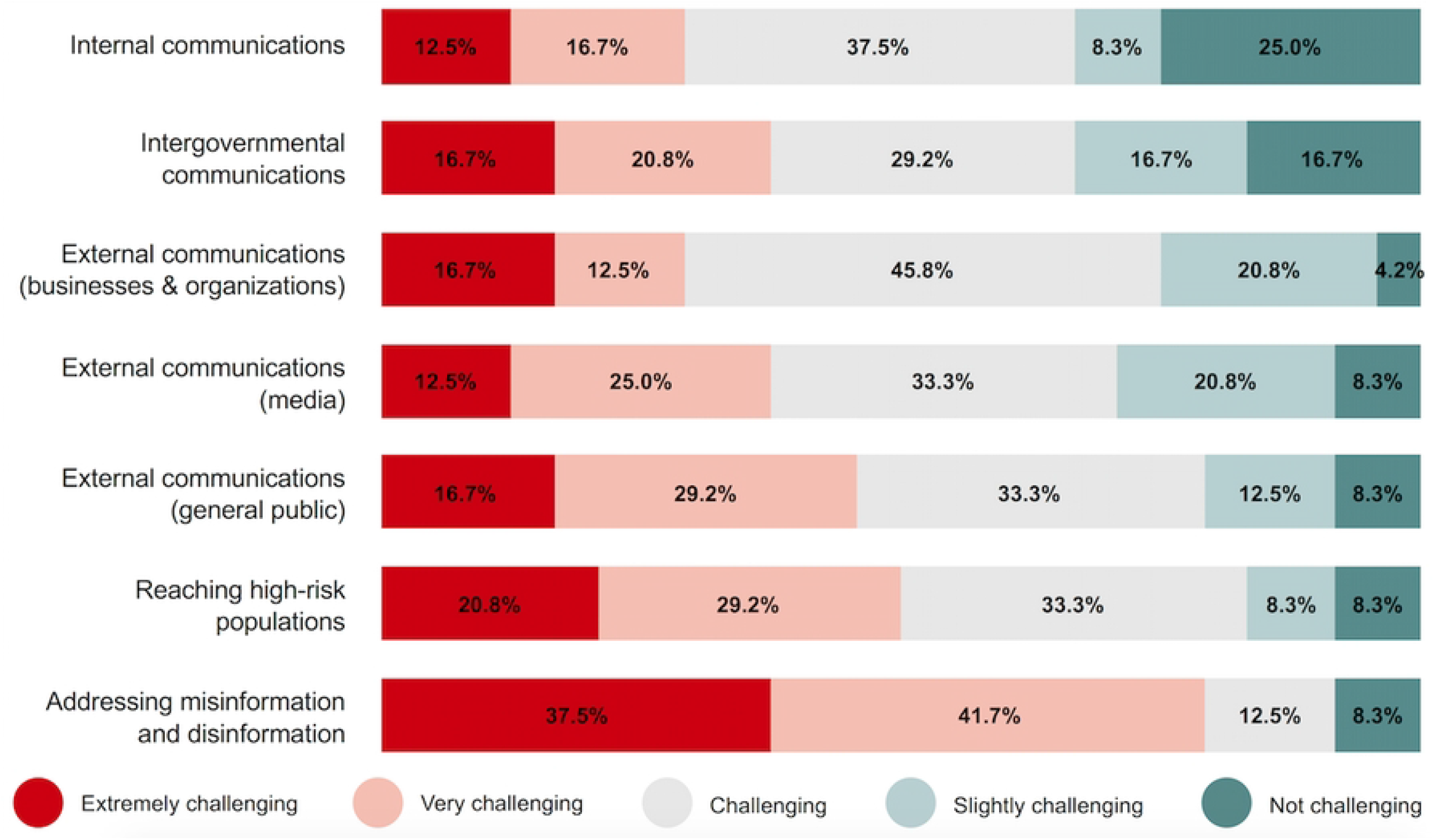
Likert-scale results regarding how challenging pandemic communication activities were perceived to be in cities (n = 24).

### Future Pandemic Preparedness – Engagement and Improvements

When asked about how their city would engage in pandemic or epidemic preparedness and response following the COVID-19 pandemic, 23 study participants (96%) reported that they anticipated their city would be more engaged and 1 participant (4%) reported that they were unsure. Local authorities reported being open to several forms of support for improving future preparedness, with 21 participants (87.5%) indicating that they anticipated their cities would be open to direct financial assistance, 15 participants (62.5%) indicating that they would be open to material assistance, and 14 participants (58.3%) indicating that they would be open to personnel assistance, and 20 participants (83.3%) indicating that they would be open to technical assistance.

When asked about how to improve future pandemic preparedness and response in their city, several key themes emerged including coordination, health systems strengthening, human resources, preparedness, resourcing, risk communication, and surveillance and information systems. The most commonly discussed themes were risk communication and surveillance. Survey responses discussing coordination included the need to improve coordination between levels of government, coordination between sectors, and coordination between local health authorities, including those in the same country as well as other countries. Human resources responses included expanding the current workforce, providing training for existing health care workers, and having teams of reserve health care workers in the case of emergencies. Preparedness responses included reviewing the legal frameworks between levels of government, passing new policy related to addressing inequities and social determinants of health, and planning for future infectious disease outbreaks. Resourcing focused responses included both sustained financing for routine public health, as well as the ability to rapidly mobilize financing and other resources in the case of an emergency. Risk communication responses included education campaigns, strategies for communicating with the media, using social media, and methods for addressing mis- and disinformation. Many of the responses focused on surveillance and information systems included the need to digitize systems, especially as a means of increasing the accessibility of data for the public and decision-makers. Survey response data are available for review in the S3 Appendix.

## Discussion

This work sought to understand how pandemic response activities in cities were coordinated with other levels of government, document when response activities were implemented, characterize how challenging response activities were, and inquire about anticipated future engagement of local authorities in pandemic preparedness and response. The results demonstrated that while national authorities were responsible and accountable for a majority of the activities considered, local governments were also responsible and accountable for key activities – especially risk communication and coordinating with community-based organizations and civil society organizations. Our results also showed that most response activities were implemented after COVID-19 had been confirmed in a city, that many pandemic response activities proved to be challenging, and that nearly all local authorities envisioned being more engaged in pandemic preparedness and response following the COVID-19 pandemic.

If it can be accepted that the COVID-19 pandemic has emphasized the importance of urban municipalities in managing complex challenges [8], this study furthers that discussion by highlighting specific activities that local governments are generally responsible for conducting and accountable for overseeing, such as risk communication and local-level coordination with community-based organizations and civil society organizations. Other key activities that local governments play large roles in, relative to other levels of government, include mandating individual behavior changes and contact tracing activities.

Effective risk communication channels are required to ensure that public health interventions, which may be highly disruptive to society, are perceived as legitimate and warranted by society [16,17]. Because of the relationship between local governments and the populations they serve – one often characterized enhanced trust compared to other governments and by an unparalleled knowledge of the local context – they are uniquely positioned to conduct risk communication activities well. Still, that is not to say that these activities did not come without their challenges throughout the pandemic response. For example addressing misinformation and disinformation was reportedly extremely or very challenging for 19 of the surveyed cities.

These challenges were not unique to the local level, however. Just three days after declaring the outbreak a Public Health Emergency of International Concern, the WHO noted that an “over-abundance of information – some accurate and some not” was complicating outbreak response and making it challenging for people to find reliable information and guidance [18].

Indeed, this problem persisted throughout the pandemic and has resulted in unique challenges for health authorities at all levels of governance [19]. Other work focused on risk communication and community engagement identified leveraging information technology, social media, and community and religious leaders as one way to effectively address rumors and misinformation [20]. Our results seem to further this position, as risk communication, including strategies focused on leveraging social media, was a frequently discussed theme for improving pandemic preparedness and response in the future.

The other pandemic response activity that local authorities were most often responsible for and accountable for overseeing was coordinating with community-based organizations and civil society organizations. The vulnerabilities of specific local communities are best assessed by the local government, which then bears the responsibility of translating that assessment into action, as appropriate [21]. In the context of public health, this may include local governments relying on established relationships to involve local organizations in addressing specific vulnerabilities and risks. Put another way, compared to higher levels of government, local governments are uniquely positioned to capitalize on their nuanced knowledge of local-level risks, vulnerabilities, and other cultural considerations to effectively engage the local community leaders and organizations as a means of supporting response efforts. Intuitively, this result is unsurprising; however, recognizing that effective community engagement is an essential component for effective pandemic response – and particularly important for reaching marginalized populations and supporting equitable responses [22] – this result holds important policy implications and future guidance for community engagement should primarily target local-level authorities.

Researchers have also discussed how data on the timing of interventions, especially at the subnational and local levels, may be important but are rarely documented [23]. As such, another important result from this work is that most response activities in cities were reactively implemented after COVID-19 had already been confirmed in their city, as opposed to more proactively implemented. A general tenet of public health response is to respond in a way commensurate with the risk to public health and to avoid unnecessary societal disruptions [24]. Accordingly, that certain measures – such as mandating business closures, mandating curfews, mandating school closures, and suspending routine services – were reactive is unsurprising given that they are highly disruptive interventions. While other activities – such as contact tracing, surging medical care, and immunization campaigns for a novel disease – are inherently reactionary in nature. Still, other activities – such as conducting data analysis and surveillance activities, risk communication, and coordinating with community-based organizations and civil society organizations – could have been conducted on a more proactive basis, especially when considering that local governments are often responsible and/or accountable for these activities.

Looking toward the future, in addition to risk communication, which was previously discussed, surveillance and information systems were a frequently referenced theme for improving pandemic preparedness and response in the future. Others have argued that the COVID-19 response accelerated previously emerging trends in urban governance, such as an emphasis on geospatial data, data-driven decision making, and app-based forms of governing in urban management [25]. For the COVID-response, these have been important for evidence-based decision making, in which data are used in real-time to inform policy decisions and interventions [8]. Our results corroborate this, as electronic and digitalized surveillance and information systems were frequently cited by study participants as one method for improving preparedness in the future.

Also of importance is that nearly all of the local authorities that participated in the survey indicated that they anticipated being more involved in pandemic and epidemic preparedness efforts following the COVID-19 pandemic. While this topic is notoriously plagued by a vicious cycle of panic and neglect, this result indicates that there is at least an appetite for more sustained engagement in the space at the local level. This may be especially true if specific financing and resources are provided or if pandemic preparedness is integrated with other urban agendas focused on health, sustainability, and resilience. Should this happen, working directly with cities to provide support – especially financial and technical, which were the indicated preferences according to survey results – could represent a practical way to improve pandemic and epidemic preparedness.

Future research agendas may also be informed by the results of this study. Of the pandemic activities considered in this study, mandating business closures, mandating individual behavior change, and financing emergency public health activities were consistently characterized as the most challenging, and could represent three key areas for future research. More specifically, efforts focused on further describing specific challenges associated with these activities may lead to identifying interventions and policies that improve future preparedness and response efforts. Relatedly, our results also suggest that research focused on best practices for using social media for risk communication and addressing mis- and disinformation during public health responses could hold important implications for improving the local response to infectious disease outbreaks. Finally, while the RACI matrices from this study are important and useful for describing the coordination structures as understood by local health authorities, future work may also wish to investigate the validity of these findings from a legal perspective (i.e., the coordination structures as outlined by legal frameworks).

These results must be interpreted in a fashion consistent with their limitations, of which there are several. For instance, surveys were completed by one local health authority per city. While we endeavored to distribute the survey to individuals who were heavily involved in the local response, and therefore would have greater knowledge about the coordination, timing, and challenges associated with pandemic response activities, this renders our results prone to personal biases. For example, in any given city, one local authority may have viewed a response activity as very challenging, while another viewed it as challenging, while another viewed it as not challenging at all. It is for this reason that we use descriptive statistics and seek to provide an overarching high-level narrative of the urban experience in responding to COVID-19. The use of descriptive statistics also helped to remedy issues of external validity and inappropriately extrapolating the results from specific cities across governance contexts.

There are also some notable limitations associated with the results regarding the timing of response activities. The attempts of local health authorities to describe when activities were implemented relative to the confirmation of COVID in various geographies are admirable.

However, their best efforts cannot overcome challenges presented by incomplete surveillance. Just as COVID-19 was not detected and reported in China until late December 2019, it is plausible that it may have emerged before that and was circulating undetected throughout populations [26]. Similarly, COVID-19 may have been circulating in a country or in a city well before it was detected and confirmed by health authorities, which could render any assessment of the relative reactivity or proactivity of response activities inaccurate. Further, many of the activities in question – mandates, risk communication, testing and contact tracing – were rapidly implemented over the span of a few days or weeks in March of 2020, unfortunately, rendering the results prone to recall bias.

In conclusion, this study showed that, local governments are often responsible and accountable for many pandemic response activities and that relative to other levels of government, local governments are most often responsible and accountable for risk communication activities and coordinating with community-based and civil society organizations. The results also suggest that most response activities were reactive, as opposed to proactive, that many response activities were perceived to be challenging, and that many local authorities envision a future in which they are more engaged in pandemic and epidemic preparedness. This type of descriptive work represents an important first step in building an evidence base for important work focused on improving the ongoing local-level response to the COVID-19 pandemic as well as future pandemics – ultimately benefiting urban and global health.

## Data Availability

All data used and/or analyzed during the current study are included in this published article and supporting information.

## Acknowledgements

Our sincere thanks to Mariana Espinosa Estrada, Charity Hung, Ramya Kancharla, Sherissa Ng, Joseph Ngamije, Grace Pickens, and Tara O’Rourke of Vital Strategies for their assistance in the distribution of survey and study materials.

## Notes

### Competing Interest Statement

The authors have declared no competing interest.

### Funding Statement

The authors received no specific funding for this work.

### Author Declarations

All study materials were submitted for ethical review to the Georgetown University Institutional Review Board, which determined that the research posed no more than minimal risk to study participants and granted an exemption to the study (STUDY00003948).

